# Current Physical Therapy for Skin Scars Management: A Scoping Review Protocol

**DOI:** 10.1101/2024.05.14.24307367

**Authors:** Sara Di Serio, Matteo Congiu, Silvia Minnucci, Valentina Scalise, Firas Mourad

## Abstract

**Background:** Scar impairments impose a great economic burden and influence a subject’s well-being and quality of life. Despite that, physiotherapy interventions are poorly investigated.

**Objective of the study:** Provide a comprehensive overview of studies addressing physiotherapy and conservative non-invasive interventions for skin scar management, summarizing studies based on scar type, localization, patient’s characteristics (e.g., age), safety and tolerance of physical interventions. The realization of an infographic will assist clinicians and patients with scars’ management. Moreover, any knowledge gaps will be identified.

**Methods:** The review will be conducted following the Joanna Briggs Institute Manual for Evidence Synthesis. MEDLINE Central, PEDro, Embase, Cochrane Library and Central Register of Controlled Trials (CENTRAL) and CINAHL and grey literature (e.g., Google Scholar) will be searched for studies considering physical therapy interventions in scars management. Every study considering conservative non-invasive physiotherapy interventions for scar management will be included. This review will look at studies carried out in any context. Articles written in English or Italian will be considered. No temporal or publication type restrictions will be placed. Selection and extraction of data will be done by three reviewers independently, any discrepancies will be resolved by a fourth reviewer. The results will be illustrated using descriptive statistics and summarized in an infographic.

**Ethics and dissemination:** No ethics approval will be necessary.

## INTRODUCTION

Scar-related impairments affect 100 million individuals every year in high-income countries^1^; of these, 11 million will develop keloids^2^. Scar occurrence and irritation is frequently attributed to trauma, burns, surgery, vaccination, skin piercing, acne, and herpes zoster. Additionally, these conditions are often accompanied by intermittent pain, persistent itching, and a feeling of constriction^3^. Under normal circumstances, immature scar undergoes a maturation process that may last several months. However, the maturation process may be limited as the inflammatory process continues within the scar. As a result, the immature stage is prolonged resulting in pathological scars such as keloids and hypertrophic scars^4^. Mechanical forces, genetics, lifestyle choices, hormonal factors like estrogen, and conditions like hypertension and pregnancy contribute to the development of pathological scars^4^. Pathological scars are mainly divided in keloids and hypertrophic scars. Hypertrophic scars have been observed to be associated with negative physical and psychological consequences such as scar contracture, limited range of motion, increased pain, itching, anxiety, and decreased quality of life. Although growing evidence highlights the impact on acceptability and the influence on quality of life, scar burden is an inadequately addressed topic and most of the studies are focused on pharmacological or surgical approaches. Recents studies on scar management observed the positive role that physical interventions may play in scar care. However, an overview of the various interventions is needed. Two previous systematic reviews^5,6^ found positive effects of massage, lotions, silicone, splinting/casting, and modalities (e.g., shockwave), however none of them included common interventions such as patient education, manual therapy or exercise. Thus, the heterogeneity among the protocols adopted in clinical trials limits the generalizability of results. By providing the most frequently used application procedures in literature, this scoping review can aid future research towards a more standardized approach.

According to the Joanna Briggs Institute (JBI), scoping reviews are used to map and clarify key concepts, identify gaps in the research knowledge base, and report on the types of evidence that address and inform practice in the field^7^. Accordingly, the present scoping review aimed to:

1. Provide a comprehensive overview of all studies addressing physiotherapy and non-invasive interventions for skin scar management, summarizing studies based on scar type, localization, patient’s characteristics (e.g. age), safety and tolerance of physical interventions.
2. Inform clinicians and patients on the topic via the realization of an infographic.
3. Identify any gap in the knowledge of physical interventions in scar management.

### Review Questions

1. What is known from the existing literature about physiotherapy and conservative non-invasive interventions in scar management?
2. Is there a relationship between the results obtained from a proposed intervention and scar type, scar localization and patient’s age?
3. What are the diagnostic procedures for scar assessment?
4. What are the current evidences regarding the safety of physical therapy modalities in scar management, in terms of adverse reactions, delayed scar maturation or deterioration of scar parameters, and patient tolerance?

## METHODS

The JBI Manual for Evidence Synthesis will be used in conducting the proposed scoping review^7^. The protocol was written using the template proposed by Lely et al.^8^. The 6-stage methodology suggested by Arksey and O’Malley^9^ will be followed: (1) identification of the research question, (2) identification of relevant studies, (3) selection of studies, (4) charting of data, (5) summary and report of results. The final step (6 optional consultation process) will be not included as considered unnecessary in the context of the current study. The Preferred Reporting Items for Systematic reviews and Meta-Analyses extension for Scoping Reviews (PRISMA-ScR) Checklist for reporting will be used to report the final manuscript^10^. We will follow the framework of Population, Concept and Context (PCC) proposed by The Joanna Briggs Institute to describe the elements of the inclusion criteria^7^.

### Eligibility Criteria

This scoping review will include systematic reviews, meta-analyses, narrative reviews, randomized controlled trials (RCTs), clinical controlled trials (CCTs), qualitative studies, case reports, case series, reviews, expert opinions, study protocols, letters, conference abstracts and grey literature. Primary sources will be excluded if already been incorporated into an included evidence synthesis unless the data they contain are not otherwise reported in the evidence synthesis. Research involving animals, *in vitro* studies and articles written in a language other than English or Italian will not be included. No temporal restrictions will be applied. The eligibility criteria are listed in the Table I.

**Table I.**

|  |  |
| --- | --- |
| **Population (P)** | "Cicatrix"[Mesh], "Cicatrix, Hypertrophic"[Mesh], "Tissue Adhesions"[Mesh], "Keloid"[Mesh], "Scar*", "Scarring", "Hypertrophic Scar*", "Burn Scar", "Contracture Scar". |
| **Concept (C)** | "Rehabilitation"[Mesh], "Exercise Therapy"[Mesh], "Physical Therapy Modalities"[Mesh], "Musculoskeletal Manipulations"[Mesh], "Therapy, Soft Tissue"[Mesh], "Exercise"[Mesh], "Self Care"[Mesh], "Patient education as topic[Mesh], "Muscle Stretching Exercises"[Mesh], "Conservative Treatment[Mesh], "Exercise", "Habilitation", "Rehabilitation", "Physiotherapy", "Physical Therapy", "Manual Therapy", "Patient Education", "Mobilization", "Resistance Training", "Strength Training", "Stretching", "Conservative Treatment", "Noninvasive Treatment", "Soft Tissue Therapy". |
| **Context (C)** | / |

#### Population

all types of scars on humans with no age restrictions will be included except for those who interfere with wound healing such as post-operative or trauma infections, those on medications that interfere with wound healing (such as steroids or chemotherapy), those with co-occurring conditions like diabetes, those with restrictive skin disorders like scleroderma or active dermatologic conditions, pregnant individuals, and those who have undergone prior scar surgery will not be included in the analysis. Additionally, acne scars will be excluded from consideration due to their different genesis and potential different outcomes^11^. The exclusion of individuals with infection, comorbidities, skin disorders and medication is justified due to their potential influence on wound healing and remodeling^12^.

#### Concept

all types of conservative non-invasive interventions including education and self-management strategies (e.g., scar massage, soft tissue mobilization, splinting…) will be included.

#### Context

Studies will be included regardless of geographical location, social or cultural context, or level of care.

### Information Sources and Search Strategy

The research strategy will be developed by four reviewers, three of whom will be directly involved in the article screening process, while one will supervise the team and address any discrepancies. To ensure rigorous and clinically relevant review outcomes, the research team comprises authors with expertise in evidence synthesis, research methodologies, as well as physiotherapists specialized in wrist and hand rehabilitation with a particular focus on scar management (from burns or post-surgery). MEDLINE Central, PEDro, Embase, Cochrane Library and Central Register of Controlled Trials (CENTRAL) and CINAHL will all be searched. At least the first 10 pages of Google Scholar will be searched for grey literature^13^. In addition the reference list of included studies will be searched manually to identify any additional studies that will be relevant. A preliminary search strategy will be conducted on PubMed in order to identify relevant keywords and terms for developing a final search strategy across all databases. The search query considers MeSH terms and free text terms, the included terms were discussed among authors. The search strategy is reported in Table I and Table II. The appendix I will report the search strategies employed for different databases. Zotero (Roy Rosenzweig Center for History and New Media Elsevier Incorporation, Fairfax, Virginia, USA) will be used for citation management. The PRISMA-S will be used to report the search strategies^14^.

**Table II.**

| Database | Query strings, keywords and boolean operators | Results |
| --- | --- | --- |
| Medline (via PubMed) | ((((((((("Scar") OR ("Scarring")) OR ("Hypertrophic Scars")) OR ("Burn Scar")) OR ("Contracture Scar")) OR ("Cicatrix"[MeSH Terms])) OR ("Cicatrix, Hypertrophic"[MeSH Terms])) OR ("Keloid"[MeSH Terms])) OR ("Tissue Adhesions"[MeSH Terms])) AND (((((((((((((((((((("Soft Tissue Therapy") OR ("Rehabilitation")) OR ("Habilitation")) OR ("Physiotherapy")) OR ("Physical Therapy")) OR ("Exercise")) OR ("Stretching")) OR ("Resistance Training")) OR ("Strength Training")) OR ("Manual Therapy")) OR ("Mobilization")) OR ("Patient Education")) OR ("Conservative Treatment")) | 3223 |

### Study Selection

The articles will be screened for inclusion using the eligibility criteria. Firstly, three reviewers will independently screen all titles and abstracts to select eligible articles, then full-text screening will be performed. In particular, for both screening phases (title and abstract, full-text), the first reviewer will screen all articles, while a second and third reviewer will screen a random percentage of articles. Articles lacking an abstract will automatically proceed to the full-text review phase. If full-text articles cannot be retrieved, authors will be contacted with a maximum of two attempts on a weekly basis. Discrepancies among reviewers will be discussed, and if consensus is not reached, a fourth reviewer will be consulted. The web-based software platform Rayyan (Rayyan Systems Incorporation, Cambridge, USA) will be used for the selection process and duplicate record removal^15^. The screening schedule will consider a minimum of 400 articles per week. Regular team meetings will take place to improve understanding of project’s objectives, discuss discrepancies and provide updates on the review process with regards to predetermined deadlines. Accounting for new potentially relevant terms, concepts, and even locations of evidence, search and eligibility criteria may be modified during the process. If changes occur, they will be reported in the appendix and registered protocol will be updated. The appendix will include a report on all excluded sources along with an explanation for their exclusion.

### Data Extraction, Synthesis and Presentation of Results

The first reviewer will create the data charting form following the JBI Manual for Evidence Synthesis for Scoping Reviews, and it will be finalized in a meeting with the review team^7^. Data will be independently cleaned by three reviewers in the same modality described for “study selection” inserting relevant data in a Microsoft Excel file (Microsoft Corporation, Redmond, WA, USA) which will be included in the appendix. PRISMA flow diagram will be used to graphically depict the flow of information through the review process^13^. The data extraction rules will be detailed in the data extraction form, Table III reports the model that will be used. The information gathered will take into account the following: (1) study design, authors and year of publication; (2) objective of the study; (3) patient population characteristics, such as sample size, demographic details, scar types and localization; (4) concept related to intervention parameters, such as type of intervention, duration, and frequency also including feasibility, safety and accessibility; (5) context such as location of care and geographical location of care; (6) outcomes, relevant results and considerations. Extracted data will be divided by kind of intervention in relation to the study question examining the effects of physical therapy on scar treatment. Discrepancies between reviewers will be discussed and if consensus is not reached, a fourth reviewer will be consulted. In case of missing data, authors will be contacted with a maximum of two email attempts on weekly basis. If no response is received, the variable will be identified as “Not Reported” if any information is lacking, and as “Unclear” if any data is conflicting or incomplete. Whether there will be several publications released from the same study (referred to as “friend studies”), only the study with the largest sample size will be taken into account^16^. Data extraction schedule plans to extract relevant data from a minimum of 50 articles per week. Regular meetings will take place regularly to identify any problems and discuss updates to the review process with pre-established time frames. Any modifications to the data extraction strategy will be reported in the results section of the final ScR.

**Table III.**
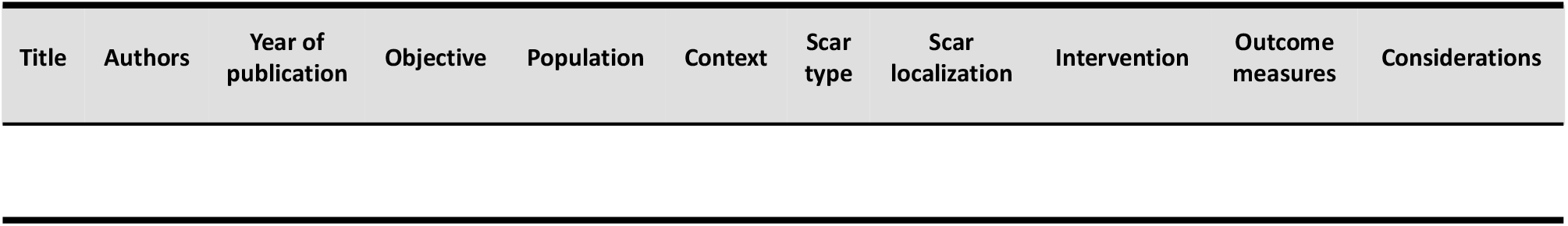

Data presentation will use descriptive statistics and will be summarized through tables and diagrams if required. At First, the types of interventions present in literature will be discussed along with the rationale and objective for their use and outcomes recorded. Then, a summary of the existing literature about the effects of therapeutic interventions in relation to scar’s characteristics, localization and subject’s age (i.e., neonats till childhood 0-12 years, adolescents 13-18 years, adults 19-44 years, middle-aged 45-64 years, aged 65+ years) will be realized. The results for every variable will be arranged in tables that include the research population, sample size, intervention type, duration, frequency, adverse effects and tolerance, as well as the major conclusions from this scoping review. The findings from this scoping review will be condensed and visually represented in an infographic.

The method used for collecting and presenting the findings may alter as the review process progresses; if there is a modification, it will be reported in the registered protocol. No additional advisors or collaborators are allowed to take part in the findings synthesis at this time of protocol registration.

## Data Availability

All data produced in the present study are available upon reasonable request to the authors.
All data produced in the present work are contained in the manuscript.

## ETHICS AND DISSEMINATION

This scoping review does not require ethics approval. An infographic summarizing the findings from the given study will be shared with physical therapists and patients in order to provide relevant information regarding therapeutic interventions for scars, patient education and self-management. The results are intended to be published in peer-reviewed journals. The researchers also intend to form recommendations for areas of future research.

## CONTRIBUTORS

The review project was initiated by Sara Di Serio, who also defined the framework. All authors designed the protocol, reviewed the manuscript and approved the final version.

## ACKNOWLEDGEMENTS

Nothing to report.

